# Oral aspirin for preventing colorectal adenoma recurrence: a systematic review and network meta-analysis of randomized controlled trials

**DOI:** 10.1101/2022.12.15.22283547

**Authors:** Khanh Dinh Hoang, Jin-Hua Chen, Tsai-Wei Huang, Yi-No Kang, Chiehfeng Chen

## Abstract

Colorectal adenomas have the potential of malignant transformation if left untreated. Multiple randomized controlled trials have been performed to evaluate the efficacy of aspirin in preventing colorectal adenoma recurrence in a population with a history of colorectal adenoma but not colorectal cancer, however, the relationship between aspirin dose and colorectal adenoma recurrence remains unclear. We conducted pairwise meta-analysis, meta-regression, trial sequential analysis, and network meta-analysis of all eligible studies. The ROB 2.0 tool was used to assess the risk of bias in the studies. The confidence in network meta-analysis (CINeMA) approach was used to evaluate the confidence of the network meta-analysis results. The network meta-analysis included eight RCTs (nine reports), comprising four on aspirin (low or high dose) alone and four on aspirin combined with another medication, all compared with placebo. In the network meta-analysis, low-dose aspirin (LDA <300 mg per day) was more effective than high-dose aspirin (HDA ≥300 mg per day) and placebo, with risk ratios of 0.76 (95% CI: 0.58 to 0.99) and 0.7 (95% CI: 0.54 to 0.91), respectively. LDA was the optimal treatment relative to HDA and placebo (P-score = 0.99). In the trial sequential analysis, LDA was only more effective than placebo when the number of included participants exceeded the optimal information size; this was not the case for HDA. LDA has statistically significant efficacy for colorectal adenoma prevention, but compared with HDA, its efficacy remains uncertain. Further trials are therefore required.

## Introduction

Colorectal adenomas are common precancerous neoplasms in the gastrointestinal tract, with an annual incidence rate of 7.2%, whereas the recurrence rate of nonadvanced and advanced adenomas was 19.3% and 22.9%, respectively according to a study in Japan in 2004.(1) Colorectal adenomas are polypoid lesions that develop in the colorectal lumen either sporadically (most cases) or as a part of adenoma-related genetic syndromes.(2) If left untreated, these precursor lesions may progress to invasive carcinoma through three main molecular mechanisms: chromosomal instability, microsatellite instability, and CpG island methylator phenotype methylation.(3) In Europe in 2006, invasive carcinoma was second to breast cancer as the leading cause of death, with 412,900 cases (equivalent to an incidence rate of 12.9%).(4) Generally, the progression rate from adenoma to invasive cancer depends on the histological subtype: 4.8%, 22.5%, and 40.7% for tubular adenoma, tubulovillous adenoma, and villous adenoma, respectively.(5) In addition, an adenoma transforms into invasive carcinoma between 4 and 48 years, depending on the polyp size and histologic morphology; adenomas or other polypoid lesions can be detected and removed easily through colonoscopy.(6) However, during colonoscopy, patients may experience unpleasant sensations (mainly varying degrees of pain); thus, approximately half the patients are unwilling to return for a surveillance procedure within the following 5 years.(7)

Numerous studies have been conducted to develop methods to prevent colorectal adenomas without performing periodic colonoscopy, such as changing eating habits to include a high-fiber diet(8) or high dry-bean intake(9) or chemoprevention using multiple medications, including aspirin (acetylsalicylic acid) and other nonsteroidal anti-inflammatory drugs (e.g., celecoxib or rofecoxib).(10) Some randomized controlled trials (RCTs) have indicated that low-dose aspirin (LDA, 81–160 mg per day) is more effective than high-dose aspirin (HDA, 300–325 mg per day) or selective cyclooxygenase enzyme (COX-2) inhibitors (e.g., 400 mg of celecoxib per day).(10) Researchers have demonstrated that daily oral aspirin use can prevent colorectal tumors. For example, platelet aggregation and activation at inflammatory foci in the colorectal mucosa lead to the release of physiologically active lipids such as prostaplandin E2 (PGE2) as well as growth factors such as platelet-derived growth factors. These substances can promote the growth of stromal cells and induce COX-2 expression on colorectal epithelial cells, leading to increased PGE2 production. Consequently, epithelial hyperplasia, epithelial–mesenchymal transformation, and transformation into a neoplasm occur.(11, 12) Furthermore, aspirin and salicylic acid, which are not absorbed by the intestinal epithelium, are converted into intermediate byproducts such as 2,3-dihydroxybenzoic acid (2,3-DHBA) or 2,5 DHBA. These substances inhibit the activity of cyclin-dependent kinases (a key family of enzymes in cell cycle regulation) and other proteins; thus, they can delay cell proliferation, thereby facilitating the repair of damaged DNA, maintaining the stability of genetic material, and creating favorable conditions for immune cells to recognize tumor cells and destroy them.(13) However, most studies have not clarified the relationship between the aspirin dose and colorectal adenoma recurrence.

Therefore, the objective of this study is to clarify the relationship between the aspirin dose and colorectal adenoma recurrence and to determine the preferred aspirin dose range for clinical use for colorectal adenoma prevention.

## Methods

This systematic review and network meta-analysis was conducted in accordance with the Preferred Reporting Items for Systematic Reviews and Meta-Analyses 2015 checklist.(14)

### Eligibility criteria

#### Inclusion criteria

We only included RCTs in which patients had a history of colorectal adenoma and had no contraindications for aspirin use, such as gastric bleeding or ulcer, hemophilia, or kidney dysfunction. Before each trial was initiated, participants had undergone colonoscopy to remove all observed polypoid lesions. Participants then received either aspirin or placebo on a daily basis, with a compliance of at least 70%. The adenomas included in the analysis were noninvasive lesions.

#### Exclusion criteria

We excluded all non-RCT studies (cohort, case–control, and cross-sectional studies). We also excluded RCTs with participants who did not receive aspirin or placebo on a daily basis as well as those with evaluations of biomarkers or adenoma-related risk factors and any other nonadenoma recurrence-based studies.

### Search strategy and information sources

We searched the PubMed/MEDLINE, Scopus, and Embase databases for relevant studies every week (on Friday) to check for all possible updates. The last search was conducted on December 31, 2021. PubMed Medical Subject Headings terms and keywords were used, including “colon adenoma,” “rectal adenoma,” “colorectal adenoma,” “colorectal polyp,” “colorectal tumor,” “aspirin,” and “acetylsalicylic acid.” No restriction on publication language or search time was applied to decrease the risks of reporting and publication bias.

### Study selection and data collection processes

Standardized checklists in which all the essential inclusion and exclusion criteria were listed in detail and clearly explained were used for eligible trial selection. Duplicated studies were removed using Endnote 20.(15) Two reviewers (KDH and YNK) screened all the titles and abstracts independently, followed by full-text assessment of potentially eligible trials. The two reviewers also independently collected published data used for statistical analysis. Any discrepancies between the two reviewers (KDH and YNK) were resolved through a consensus or the adjudication of a third reviewer (CC).

### Data items and subgroup analysis

#### Primary outcome

Overall colorectal adenoma recurrence was defined as the occurrence of any adenoma (any size or histologic subtype) in the colorectum after the removal of all observable lesions at the beginning of a trial at any aspirin dose and any time. Histologically, participants with low-grade or high-grade dysplasia adenoma or in-situ carcinoma were included in the analysis.

Adenoma recurrence was stratified according to aspirin dosage, namely HDA (≥300 mg per day) and LDA (<300 mg per day); the intervention time of short-term aspirin use (≤1 year) or long-term aspirin use (>1 year); and aspirin dose (six doses of 81, 100, 160, 300, 325, and 600 mg per day).

#### Secondary outcome

The secondary outcome was the mean number of recurrent adenomas. This outcome was analyzed per patient, and all patients.

The incidence of severe adverse events was determined based on the number of participants who had at least one adverse event during follow-up.

### Study risk-of-bias assessment

The risk of bias of all the included studies was assessed using the ROB 2.0 tool.(16) The assessment results of each included study were saved in the supplementary file. The entire process was performed by two reviewers (DKH and YNK) independently. Disagreements were resolved through a consensus or based on the opinion of a third reviewer (CC). The ROB assessment results are displayed in a risk-of-bias summary figure (Table 2) created using Robvis.(17) Publication and reporting bias were assessed using a funnel plot.(18)

### Effect measures and statistical analysis

#### For pairwise meta-analysis

Revman 5.4 software(19) was used for the pairwise meta-analysis of all outcomes. To avoid overestimating the effect size and misunderstanding the results, we analyzed all dichotomous outcomes through a risk ratio estimated using the Mantel–Haenszel method with a random-effects model instead of an odds ratio.(20) Continuous outcomes (mean number of recurrent adenomas) were analyzed based on the mean difference determined using the inverse variance method with a random-effects model. Between-study heterogeneity was assessed using Tau^2^, Chi^2^, and I^2^ tests; 95% confidence interval (95% CI) and the P value of 0.05 were applied to all analyses (except Chi^2^ with p value <0.10).

#### For meta-regression

We evaluated the relationship between the aspirin dose (individual doses) and dose response (the overall colorectal adenoma recurrence according to the risk ratio) using R 4.1.0 (Rstudio version 1.41717). Meta-regression was conducted using a random-effects model and both linear and nonlinear models. The model fit was also compared using analysis of variance.

#### For network meta-analysis

The confidence in network meta-analysis (CINeMA) approach was employed to assess the certainty of evidence in the network using a random-effects NMA model.(21, 22) We rated the certainty of evidence in each domain in each comparison as “no concerns,” “some concerns,” or “major concerns.” To evaluate imprecision, heterogeneity, and incoherence, we defined the clinically key effect size (using the risk ratio) as 1.25 (the range of equivalence was defined from 0.8 to 1.25). Then, we judged the level of certainty across domains using the 4-level confidence of the GRADE approach: very low, low, moderate, or high.(23)

“*A triangle of LDA, HDA, and placebo*,” manifesting the interactive relationship between three interventions, was created using NMAstudio (version 0.1),(24) which is a web application programmed in Python and linked to the netmeta package in R software. Based on the results of the network meta-analysis, we ranked the treatments under study to determine the optimal treatment in the network by using the surface under the cumulative ranking curve measure (P-score). The results are presented in the form of a heatmap.

#### For trial sequential analysis

Additionally, we performed trial sequential analysis to make decision on whether or not to conduct further clinical trials.(25) Data pooling was performed based on the DerSimonian–Laird method, with boundaries based on the O’Brien–Fleming method, by using R software.

## Results

### Search strategy

By December 31, 2021, 7,918 articles from the three databases had been identified based on a screening of titles and abstracts. After the removal of duplicated articles and screening titles and abstracts, 50 articles were included in full-text assessment to determine their eligibility. Finally, 9 studies representing 8 RCTs(26-34) were included in the quantitative analysis (Fig 1). In 1993, Gann et al.(35) conducted a large clinical trial involving more than 22,000 participants (all the participants were male physicians). However, the participants did not receive aspirin or placebo on a daily basis and not all participants had undergone colonoscopy at the start of the trial. This study was therefore excluded.

**Fig 1.**
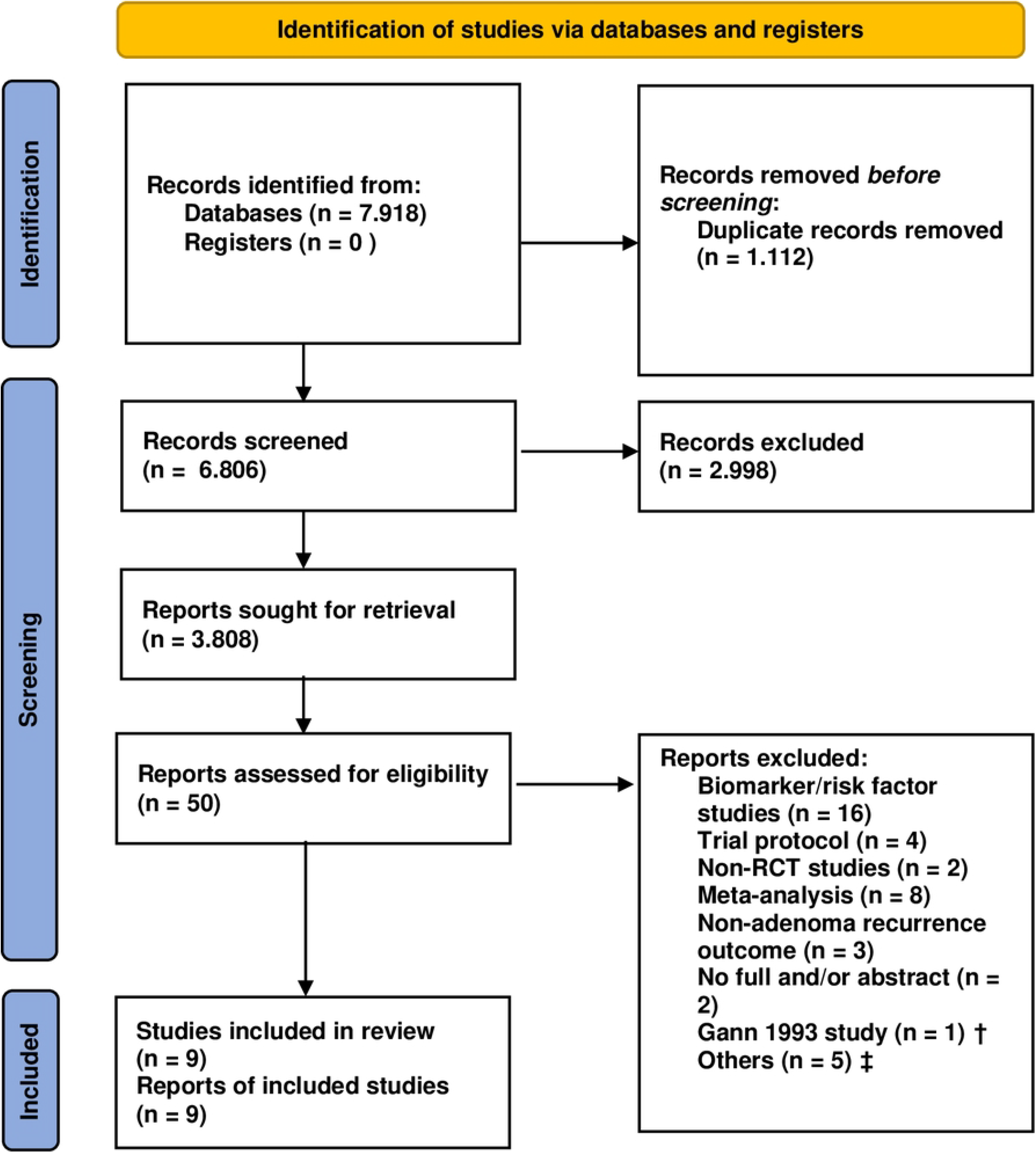
Study search flow chart. Note: † The reason to exclude this study was presented above. ‡ The reasons for others were listed in Supplementary file.

### Trial characteristics

The eight trials (nine reports) that met the inclusion criteria were published between 2003 and 2021 and involved a total of 3,011 participants. Four of the trials (five reports) compared aspirin alone (LDA or HDA or both) with placebo, and the other four compared aspirin in combination with another medication (e.g., folic acid, mesalazine, resistant starch, or eicosapentaenoic acid) with double placebo. All the study participants were randomly assigned to the intervention arms, with 1,695 participants receiving aspirin and 1,316 receiving placebo. The mean age of participants with genetic syndromes (familial adenomatous polyposis or Lynch syndrome) ranged from 30 to 59 years in three trials (Burn et al.,(31) Ishikawa et al.,(28) Ishikawa et al.,(29) and the mean age of participants from the general population was 57–69 years. The follow-up time ranged from 8 months to 5 years. Seven trials (except the study by Ishikawa,(28) which only provided data on severe adverse events) provided information on colorectal adenoma recurrence. Most trials examining the effects of different aspirin doses reported separate data for each dose, except for the study by Benamouzig et al.(33) (the data of two groups with aspirin doses of 160 and 300 mg were combined). In addition, one RCT(33) (32) was reported twice based on follow-up time (at year 1 and 4). However, for overall adenoma recurrence, we only included the study with the longest follow-up time.

**Table 1.**
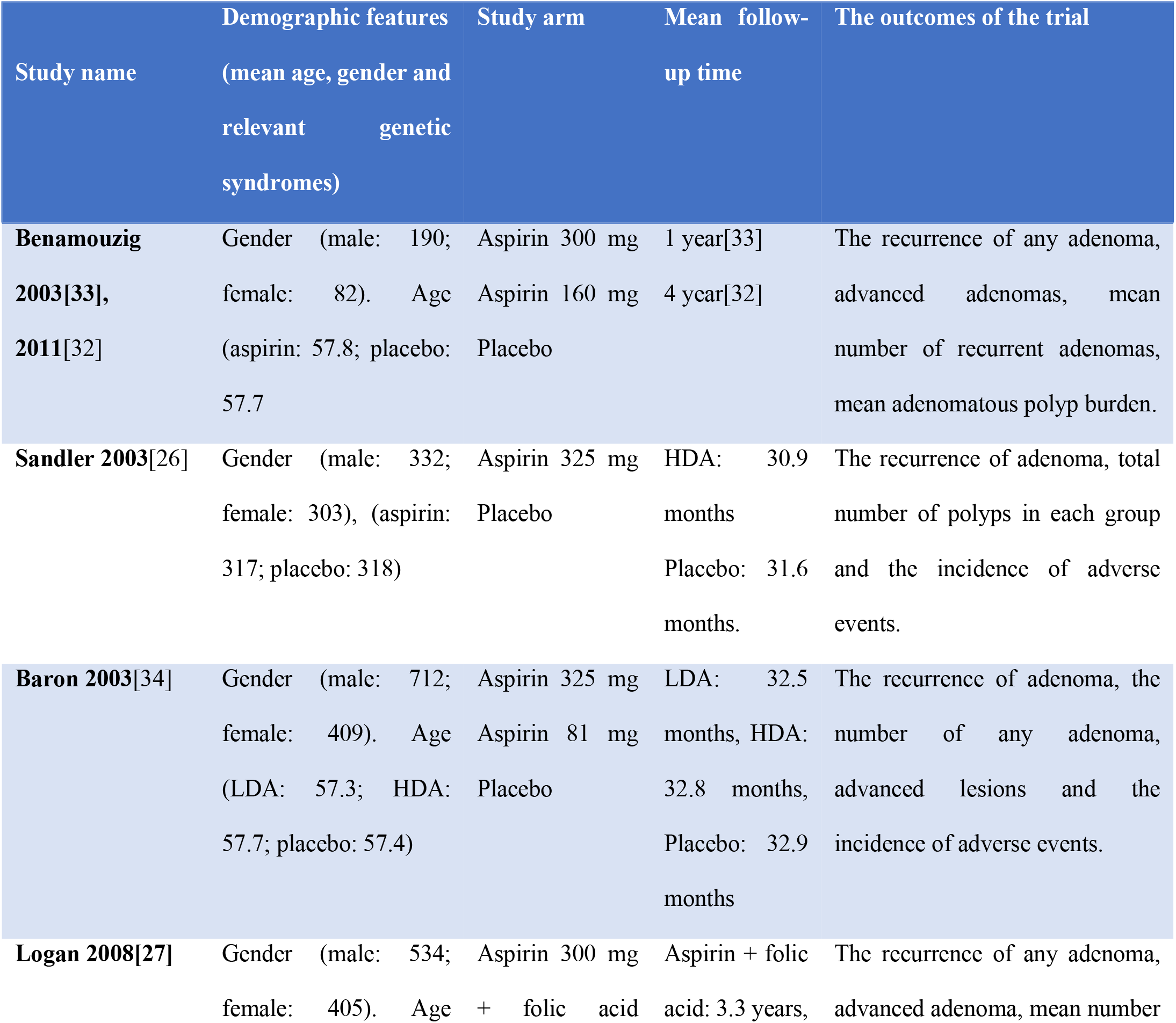
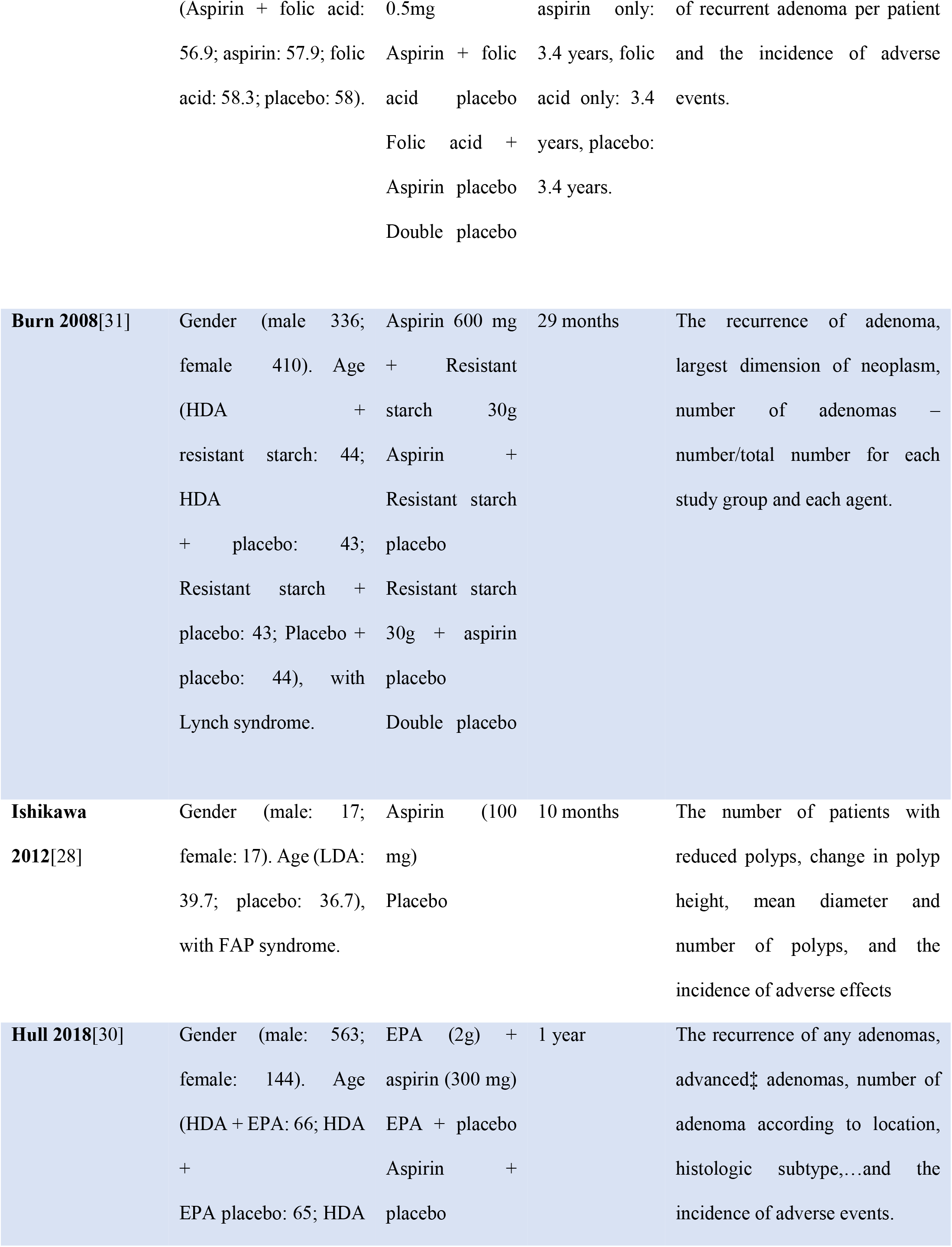
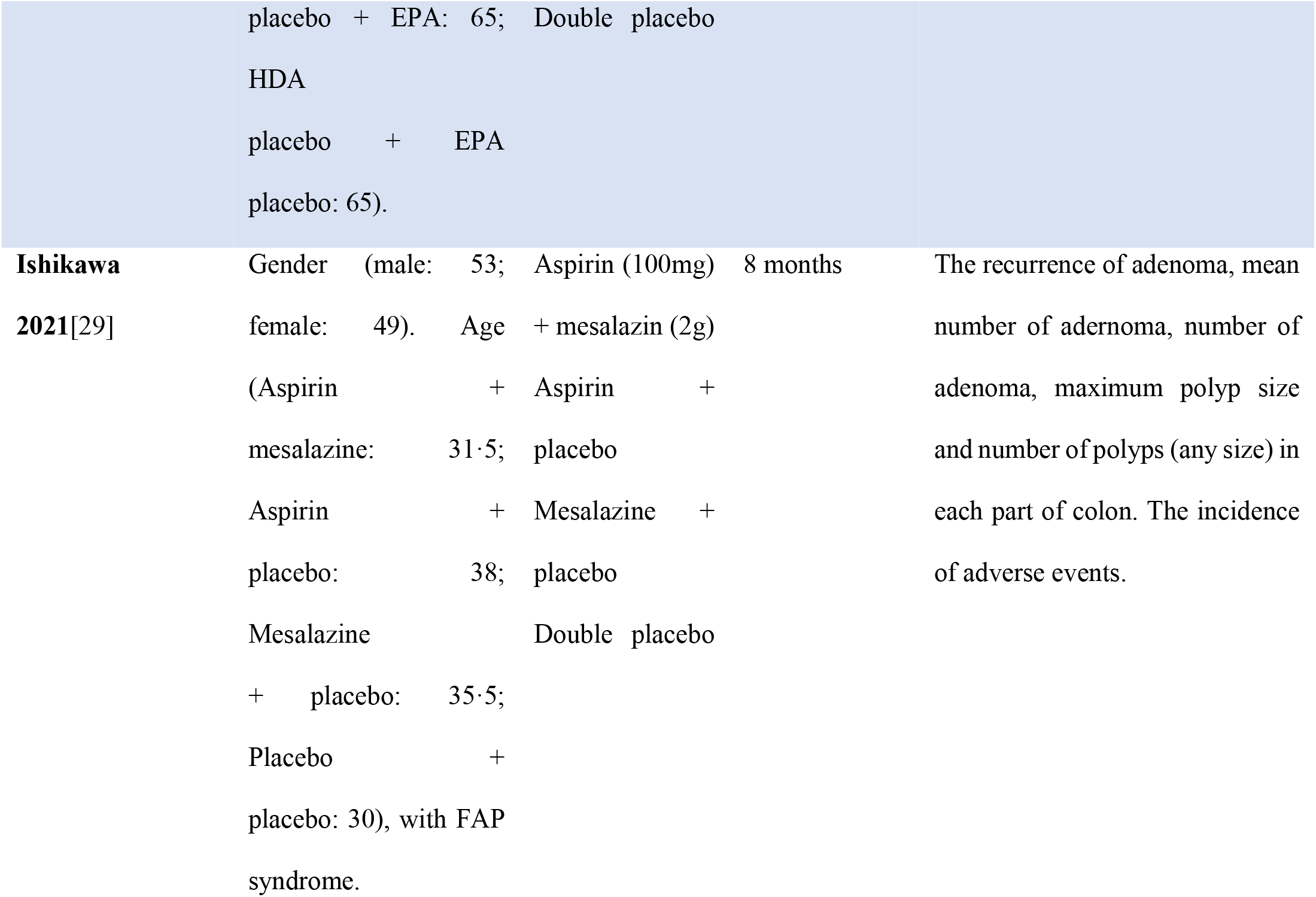
Study characteristic. LDA: Low-dose aspirin, HDA: High-dose aspirin; PLB: Placebo; FAP syndrome: Familial adenomatous polyposis syndrome, EPA: Eicosapentaenoic acid.

### Primary outcome and subgroup analysis

#### Risk-of-bias assessment (for the primary outcome)

Of seven included RCTs, four had an overall low risk of bias,(28-31, 34) one had some concerns(27) related to missing outcome data (more than 10% of participants withdrew from the trial mainly because of death, poor health status, or lack of contact with the participants), and two had an overall high risk of bias because they deviated from the intended interventions and had missing outcome data (use of inappropriate analysis and too many eligible participants were excluded from the analysis(26)) and differences between intervention groups in the proportions of missing outcome data (28.45% in the placebo group and 20.31% in the aspirin group(33) (32)). The risk of bias in the included studies is presented in S1 Table. The funnel plot for the primary outcome appeared relatively symmetrical (S1 Fig), indicating no evidence of publication bias.

#### Overall adenoma recurrence (primary outcome)

Eight reports (seven RCTs) provided data on overall adenoma recurrence.(26, 27, 29-34) In general, aspirin significantly reduced the overall recurrence of colorectal adenoma at any dose and any time during follow-up (risk ratio [RR] = 0.86, 95% CI: 0.75 to 0.99, P = 0.03, and *I*^2^ = 40%) (Fig 2). A subgroup analysis revealed no significant difference between the aspirin and placebo groups for short-term aspirin use (RR = 0.81, 95% CI: 0.59 to 1.10, P = 0.18, and *I*^2^ = 64%); however, for long-term aspirin use, a statistically significant difference was detected between the two groups (RR = 0.84, 95% CI: 0.73 to 0.98, P = 0.02, and *I*^2^ = 24%).

**Fig 2.**
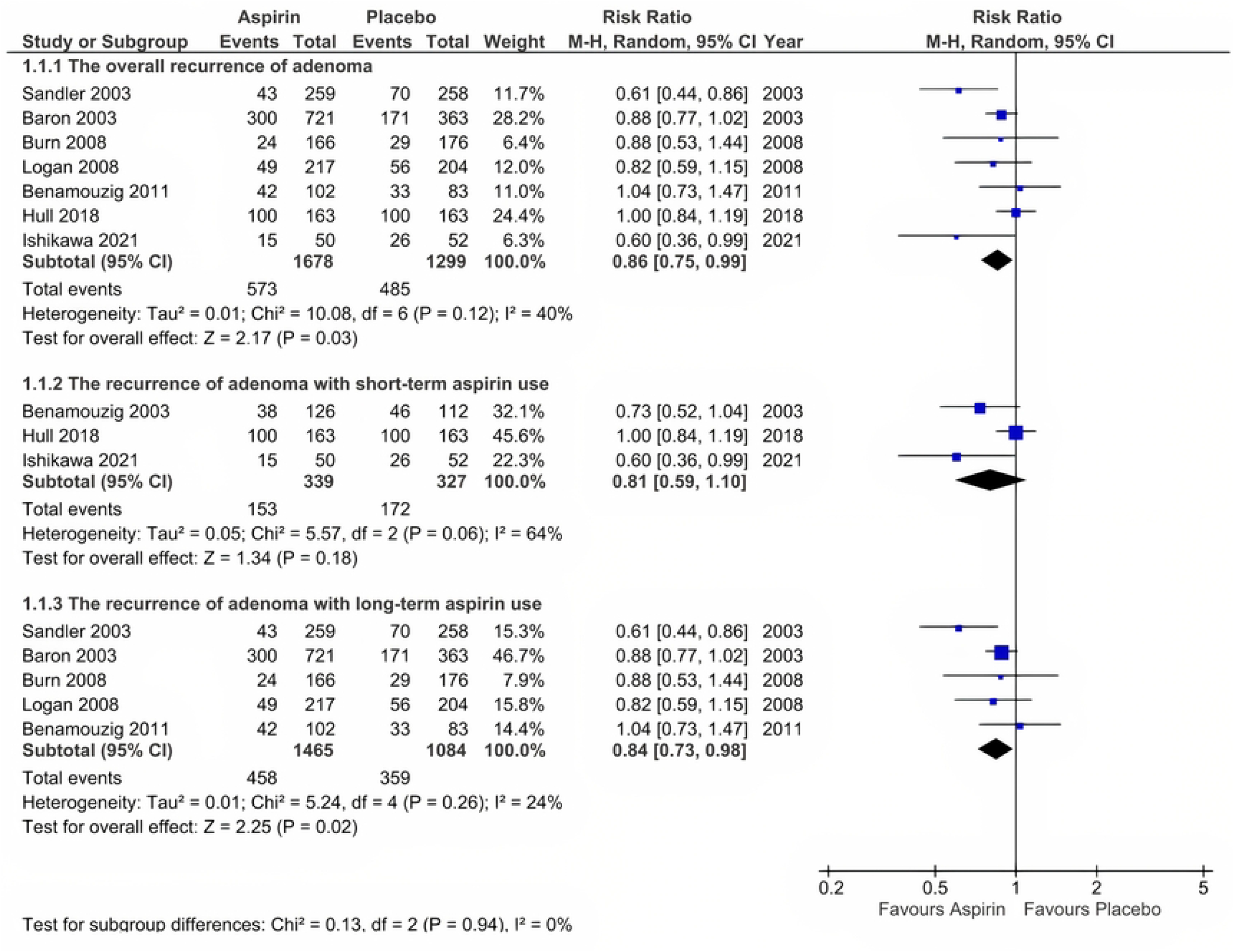
Overall adenoma recurence (the primary outcome)

#### Adenoma recurrence with low-dose aspirin use

Only three RCTs provided data on adenoma recurrence with LDA use.(29, 32, 34) Data from two reports (Benamouzig 2003 et al.,(33) and Benamouzig 2011 et al.,(32)) were based on the same trial, however, Benamouzig 2003 combined the two groups of 160 mg aspirin and 300 mg aspirin into a single aspirin group. While, Benamouzig 2011 reported into two groups “Analysis at 1 or 4 years” and “Analysis at 4 years”, which led to the inability to collect data for short-term aspirin use. At any time, LDA decreased adenoma recurrence by 22% (RR = 0.78, 95% CI: 0.67 to 0.91, P = 0.001, and *I*^2^ = 0%) (Fig 3). Moreover, the subgroup analysis revealed the same results for both short-term and long-term use of LDA (RR = 0.60, 95% CI: 0.36 to 0.99, and P = 0.05; *I*^2^ was not applicable because only one RCT was included vs. RR = 0.80, 95% CI: 0.68 to 0.94, P = 0.006, and *I*^2^ = 0%).

**Fig 3.**
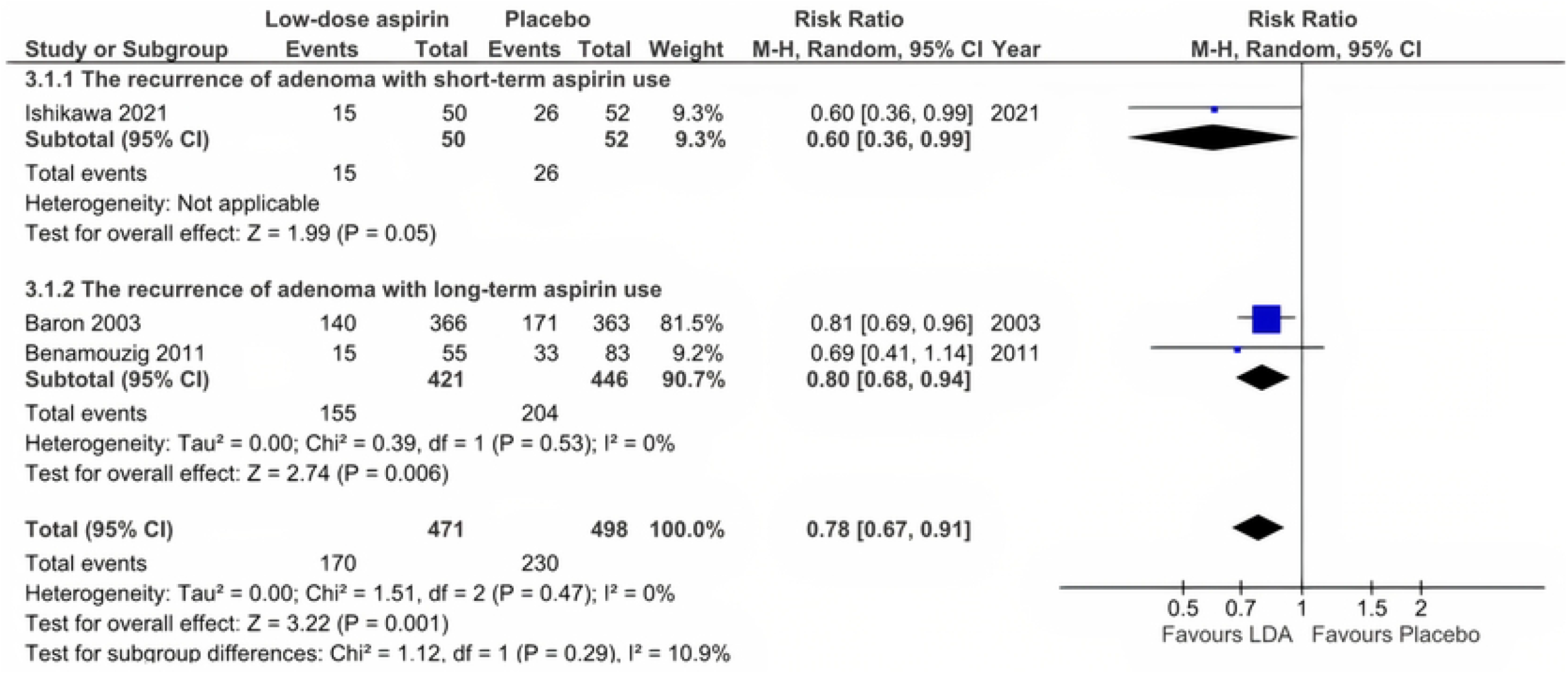
Adenoma recurrence with low-dose aspirin use.

#### Adenoma recurrence with high-dose aspirin use

Six RCTs were included in the analysis of the effect of HDA use.(26, 27, 30-32, 34) No difference was observed between the aspirin and placebo groups when participants were treated with HDA (RR = 0.93, 95% CI: 0.77 to 1.11, P = 0.43, and *I*^2^ = 62%) (Fig 4). In addition, the subgroup analysis revealed no significant difference between the aspirin and placebo groups in terms of either short-term or long-term use (RR = 1.00, 95% CI: 0.84 to 1.19, and P = 1.00; *I*^2^ was not applicable because only one trial was included vs. RR = 0.91, 95% CI: 0.71 to 1.16, P = 0.44, and *I*^2^ = 68%).

**Fig 4.**
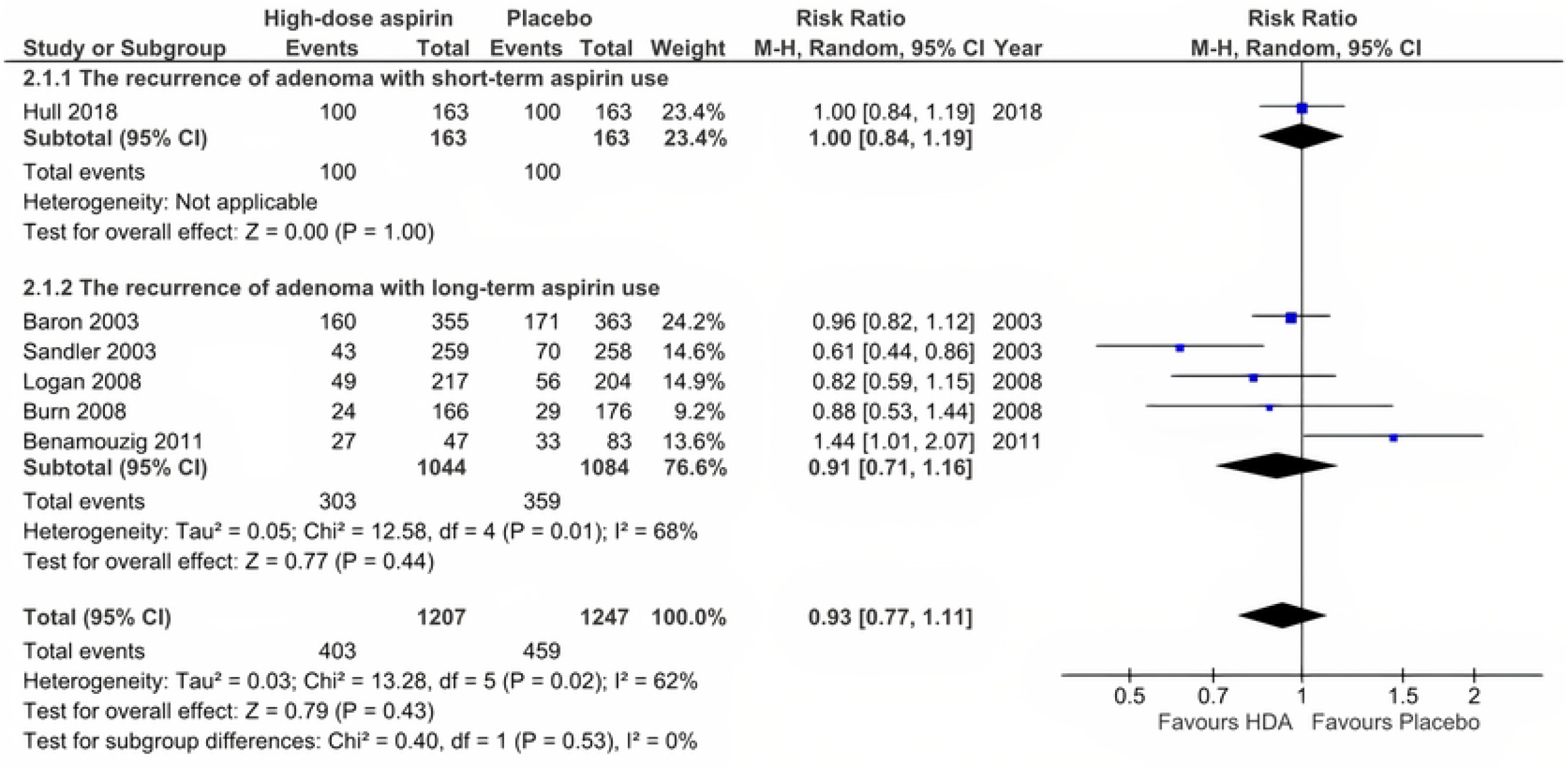
Adenoma recurrence with high-dose aspirin use.

#### Comparison of low-dose and high-dose aspirin use in relation to adenoma recurrence

For this comparison, two RCTs were included in the analysis.(32, 34) Both RCTs provided data on colorectal adenoma prevention with long-term aspirin use. One of these two trials had a high risk of bias resulting from an inappropriate analysis in which too many eligible participants were excluded from the analysis. The analysis indicated that there were no statistically significant difference between (RR = 0.67, 95% CI: 0.38 to 1.17, P = 0.16, and *I*^2^ = 79%) (Fig 5).

**Fig 5.**
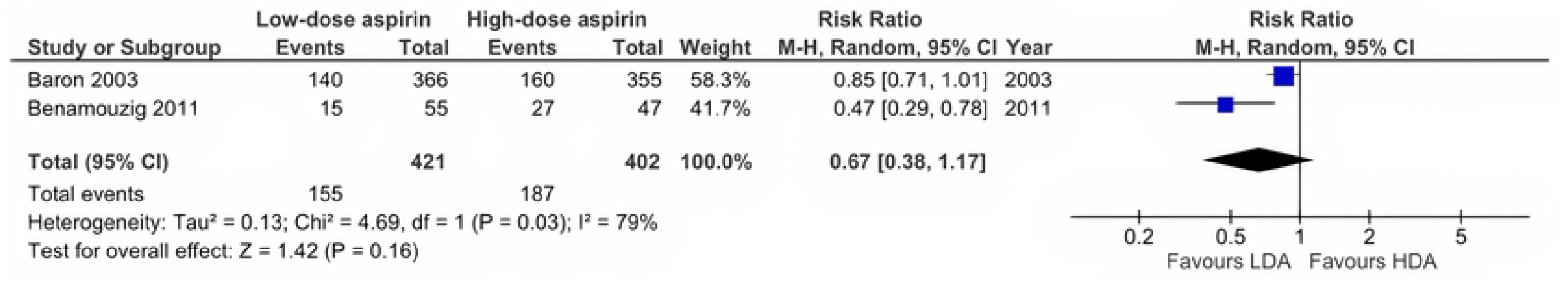
Comparison of low-dose and high-dose aspirin use in relation to adenoma recurrence. LDA: Low-dose aspirin; HDA: High-dose aspirin.

#### Sensitivity analysis for the primary outcome

After two studies with a high risk of bias (Sandler et al.(26) and Benamouzig et al.(32)) were removed from the overall adenoma recurrence outcome analysis, the results indicated that aspirin became less effective over time (RR = 0.9, 95% CI: 0.80 to 1.00, P = 0.05, and *I*^2^ = 9% vs. RR = 0.86, 95% CI: 0.75 to 0.99, and P = 0.03 for the original analysis), but this was nonsignificant. We also performed a similar analysis of the effect of LDA on colorectal adenoma prevention and identified no significant difference between aspirin and placebo groups and an increased inconsistency between studies (RR = 0.77, 95% CI: 0.61 to 0.97, P = 0.02, and *I*^2^ = 20% vs. RR = 0.78, 95% CI: 0.67 to 0.91, P = 0.001, and *I*^2^ = 0%).

### Secondary outcome

#### Mean number of recurrent adenomas

Data from six reports representing five RCTs were pooled for this outcome analysis.(26, 27, 29, 30, 32, 33) The analysis favored aspirin over placebo (mean difference −0.16, 95% CI: −0.26 to −0.06, P = 0.001, and *I*^2^ = 0%) (S2 Fig).

#### Incidence of severe adverse events

Data from six RCTs were pooled for this analysis.(26, 28, 30-32, 34) The analysis revealed that more severe adverse events were observed in the placebo group than in the aspirin group (RR = 1.29, 95% CI: 1.05 to 1.59, P = 0.02, and *I*^2^ = 0%), indicating that aspirin may be an unsafe drug for the prevention of adenoma recurrence, especially LDA (S3 Fig).

#### Sensitivity analysis for incidence of severe adverse events

For severe adverse events, of six RCTs(26, 28, 30-32, 34) included in the analysis, the study by Baron et al.(34) reported a significantly higher rate of severe adverse events in both the aspirin and placebo groups than other studies (data shown in S3 Fig). After this study was excluded from the analysis, no statistically significant difference in the incidence of severe adverse events in either the aspirin or placebo group was detected (RR = 0.99, 95% CI: 0.69, 1.43), indicating that aspirin might be a safe drug for use at both low and high doses for preventing colorectal adenomas.

### Meta-regression

Data for the meta-regression analysis were extracted from seven RCTs.(26, 27, 29-32, 34) According to the linear line (red dotted line), the most effective aspirin dose fluctuated by approximately 100 mg (RR 0.6, 95% CI: 0.36 to 0.99), and a downward trend in the effectiveness of aspirin with the increasing aspirin dose was noted. Furthermore, an analogous predisposition was observed in the left branch of the blue parabola. However, the relationship was not statistically significant in either linear or nonlinear models (P > 0.05) (S4 Fig).

### Network meta-analysis to compare low-dose and high-dose aspirin use and placebo

Data from seven RCTs were pooled for this analysis.(26, 27, 29-32, 34) The analysis revealed that LDA was favored over HDA and PLB with statistcally significant difference (NMA estimate: RR = 1.32, 95% CI: 1.01 to 1.73, and RR = 0.70, 95% CI: 0.54 to 0.91, respectively shown in League table (S5 Fig)), with moderate certainty of evidence for LDA vs PLB comparison due to some concerns in heterogeineity, but with low certainty of evidence for LDA vs HDA comparison due to major concerns in heterogeneity (shown in S6 Fig); finally, no substantial difference were observed between HDA and PLB (RR = 0.92, 95% CI: 0.77 to 1.10) with low level of certainty due to within-study bias, imprecision, and heterogeneity. The network plot of low-dose, high-dose aspirin and placebo was shown in Fig 6.

**Fig 6.**
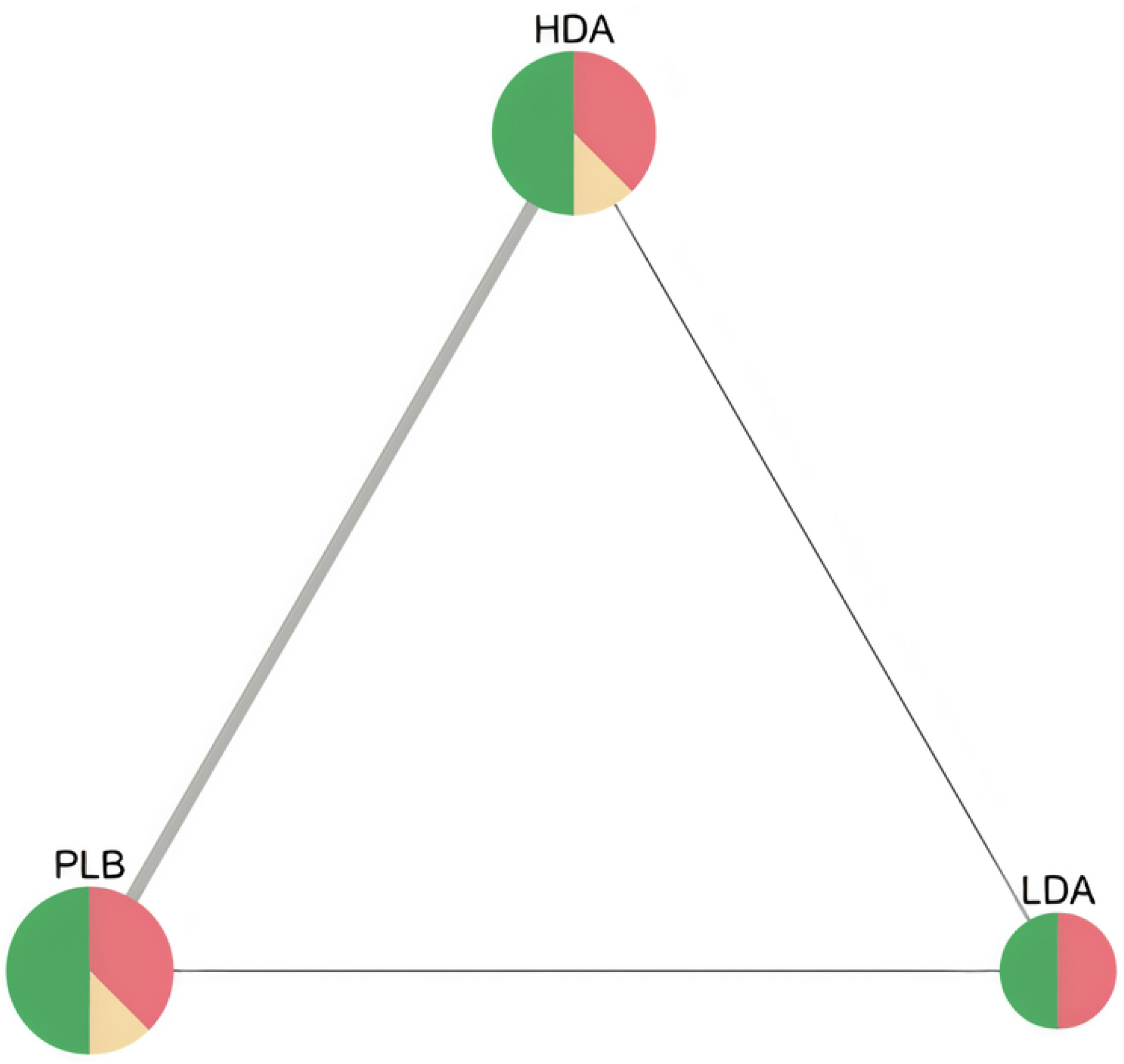
Network plot of low-dose, high-dose aspirin and placebo in RCTs for recurrence of colorectal adenomas. Note that node size is proportional to sample size, edge width is proportional to number of studies, node colour refers to majority ROB. (HDA: high-dose aspirin, LDA: low-dose aspirin, PLB: placebo).

In the P-score heatmap, the treatments (LDA, HDA, and placebo) were classified by colour from most effective to least effective (from yellow to violet), with a P-score ranging from 0 to 1, which can be interpreted as an average degree of certainty for a treatment to be more effective than other treatments in the network. The heatmap suggests that compared with HDA and placebo, LDA is the most effective treatment in the network with P-score = 0.99, whereas P-score = 0.42 for HDA and P-score = 0.1 for PLB (Fig 7).

**Fig 7.**
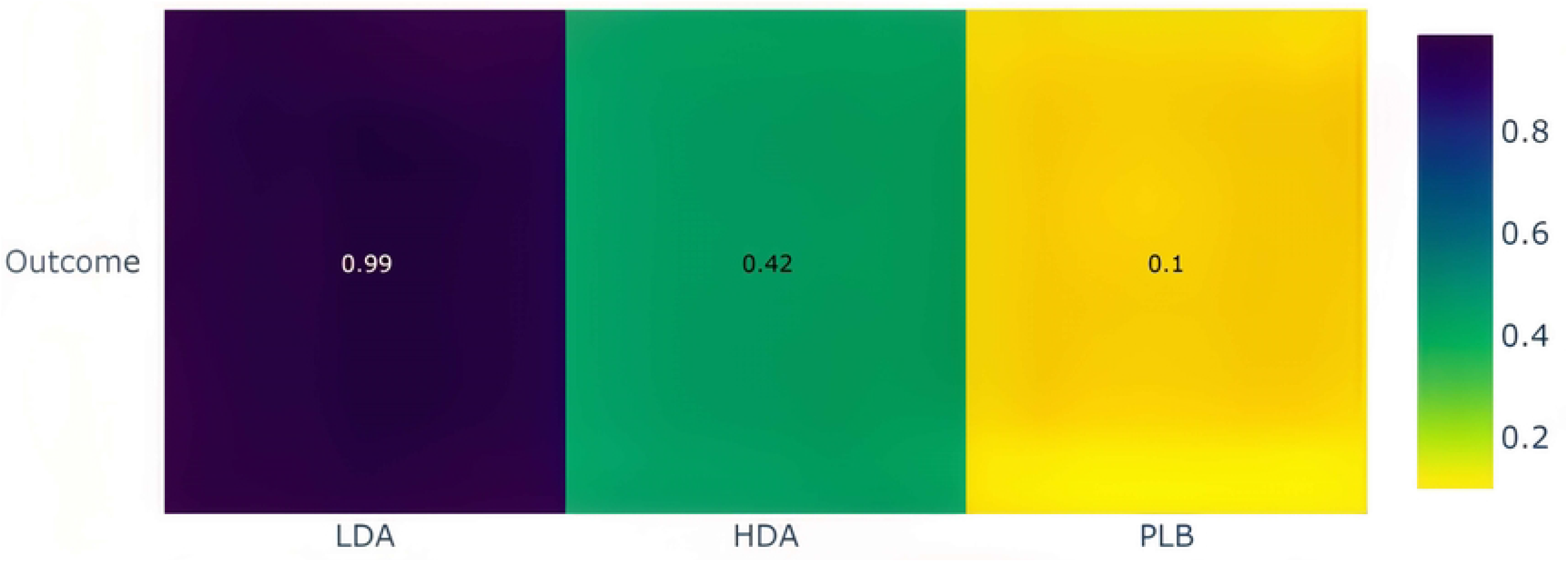
The heatmap of treatment ranking of the netwwork. Note that LDA: low-dose aspirin, HDA: high-dose aspirin, PLB: placebo. P-score ranges from 0 to 1 (equivalent to color ranging from yellow to violet).

### Trial sequential analysis

For the LDA versus placebo comparison, the last point of the Z-curve lying outside the monitoring boundary favored LDA, and the number of participants included in the analysis exceeded the required information size, indicating that the difference between the two groups was significant, and that no further trials were required (Fig 8). By contrast, no significant difference between the HDA and placebo groups or between the HDA and LDA groups was recorded in this analysis (S13 and S14 Figs).

**Fig. 8.**
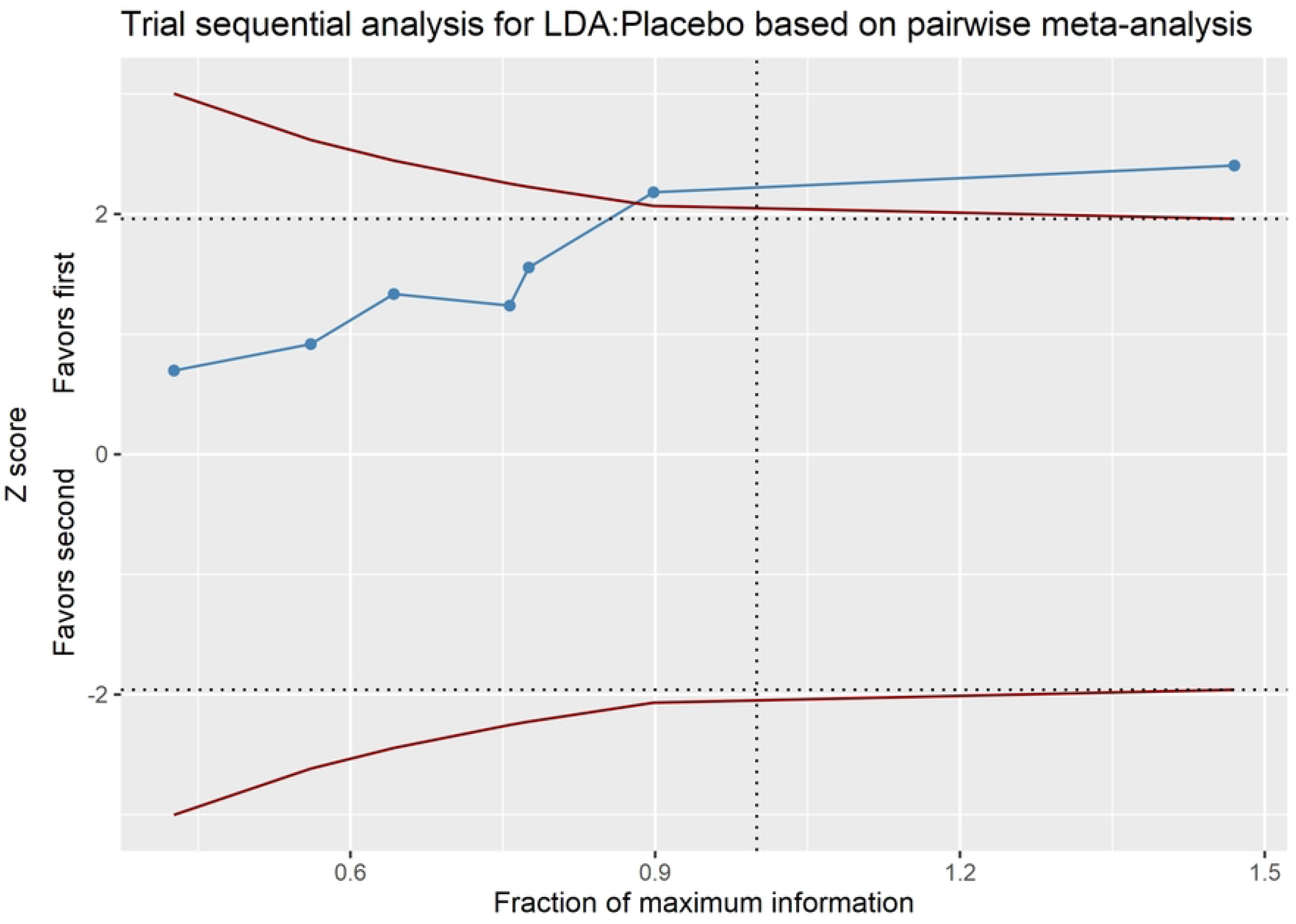
Trial sequential analysis (TSA) Note that blue line, red lines, horizontal dotted lines, vertical dotted lines represent Z-curve, monitoring boundary, required information size, conventional boundary, respectively. The areas outside the red lines indicate the area favoring LDA or placebo. LDA: Low-dose aspirin.

## Discussion

Overall, we revealed that LDA was effective for colorectal adenoma recurrence prevention, and its efficacy was more statistically significant for long-term use than for short-term use.

From the analyses, we demonstrated that aspirin use (any dose compared with placebo) reduced colorectal adenoma recurrence by approximately 14%, and the long-term use of aspirin produced a statistically more significant difference than short-term use. In addition, not only did aspirin reduce adenoma recurrence but it also helped reduce the number of recurrent adenomas after the intervention, and the difference was more pronounced with long-term aspirin use than with short-term use.

LDA was confirmed to be more effective than HDA, as demonstrated by network meta-analyses. Moreover, LDA was ranked as the most effective treatment in the network compared with HDA and placebo (as illustrated in the heatmap in Fig 8). We can therefore conclude that the long-term use of LDA is more effective for the prevention of colorectal adenoma than HDA. Although the role of LDA in adenoma recurrence was verified by Veettil et al.,(10) the authors compared LDA and HDA indirectly through the mediating effect of placebo.

However, the present trial sequential analysis indicated that LDA was more effective than placebo, and the optimal information size for the analysis was reached (Fig 6). However, the comparison of HDA versus placebo and LDA versus HDA revealed no statistical significance (S13 and S14 Figs). This seemed to be inconsistent with the results of the network meta-analysis. Furthermore, the number of participants in the LDA versus HDA comparison exceeded the required information size, whereas the sample size in the HDA versus placebo comparison remained small.

In the meta-regression, we revealed that LDA (especially LDA of approximately 100 mg) might be the optimal treatment, and the effectiveness of aspirin showed a downward trend with the increasing dose. Although we were unable to clearly explain this trend, we believe that this result has value in clinical practice. One recent study favored the use of aspirin at a dose of ≤100 mg for preventing advanced lesions.(36) Moreover, within a safe dose range, a lower dose usually provides more benefits, such as lower costs (economic effectiveness), fewer side effects, and more improved therapeutic efficacy, than higher doses.(37) However, the authors did not indicate a threshold of effectiveness for clinical use (a dose that is too low might not have any benefits), which may lead to difficulties when selecting a specific dose in practice.

In terms of severe adverse events, Baron et al.,(34) who conducted a large trial with a robust research design, identified a very high rate of severe adverse events in the aspirin group compared with those in other studies (some favored aspirin and some placebo but with a nonsignificant difference), suggesting that aspirin is an unsafe drug (RR = 1.29, 95% CI: 1.05 to 1.59). These results are not easy to explain. However, after this study was removed from the analysis, the difference in severe adverse events between the two groups became nonsignificant, indicating that aspirin might be a safe medication for clinical use (RR = 0.99, 95% CI: 0.69, 1.43).

Despite the addition of a few new trials and the provision of a new approach for robust statistical analyses, our study still has some limitations. The number of included studies was quite small, and no multiple direct comparisons between HDA and LDA were provided. Only two RCTs compared doses, and these were placebo mediated, which might reduce the reliability of the results, and the sample sizes of some studies were relatively small, with heterogeneous intervention times. In some studies, a high risk of bias may reduce the reliability of the results. This study also only evaluated the effectiveness of aspirin for up to 5 years rather than over a longer period. Finally, only a few doses were included in the design, and we did not evaluate other doses.

Compared with placebo, LDA significantly reduces the recurrence of colorectal adenomas in participants with and without adenoma-related syndromes. However, its efficacy compared with HDA remains uncertain. Further RCTs are therefore required to verify these results.

## Data Availability

All relevant data are within the manuscript and its Supporting Information files.

## Acknowledgments

This manuscript was edited by Wallace Academic Editing.

## Author contributions

KDH contributed to the research idea, study design, study selection, data extraction, statistical analysis and result interpretation, risk-of-bias assessment, and writing and editing of the manuscript; CC – Corresponding author who is acting as the submission’s guarantor contributed to the research idea, study design, statistical analysis and interpretation, risk-of-bias assessment, and editing of the manuscript; YNK contributed to the study selection, data extraction, statistical analysis and interpretation, and editing of the manuscript; TWH and JHC contributed to the statistical analysis and interpretation; all the authors approved the final manuscript.

## Data availability

Please contact the corresponding author for the data set.

## Ethical approval

Not required.

